# The equivalence of hydration status measured by the Fresenius BCM and the Cella Bioimpedance Spectroscopy devices on hemodialysis patients

**DOI:** 10.1101/2021.09.13.21263290

**Authors:** Jim Matthie, Borut Baricevic, Vlasta Malnaric Marentic, Boris Krajacic

**Affiliations:** Matthie Consulting, La Jolla, CA, USA; Borut Baricevic, Cella Medical, Inc., La Jolla, CA, USA; Novo Mesto General Hospital, Department of Nephrology and Dialysis, Novo Mesto, Slovenia; Novo Mesto Diagnostic Centre Bled, Department of Cardiology, Novo Mesto, Slovenia

**Keywords:** Hemodialysis, Overhydration, Fluid overload, Bioimpedance Spectroscopy, Fresenius BCM, Body Composition Monitor

## Abstract

**Background:** Fluid management is a serious challenge for patients undergoing hemodialysis therapy (HD). Bioimpedance spectroscopy (BIS) is a promising technique to help with clinical hydration (HYD) assessment. The Fresenius Medical Care (FMC) Body Composition Monitor (BCM) is the standard but is large and expensive. Cella Medical has introduced a small wireless BIS device. This study compared the HYD status predicted by the two devices.

**Methods:** Following the FMC BCM device manual guidelines, measurements of BIS were made wrist-ankle using typical ECG electrodes on the non-fistula side of HD patients pre dialysis while in their normal supine position. As usual, patients were measured before their normal time of therapy with the BCM. The Cella measurements were then performed within two minutes.

**Results:** Forty-two HD patients (M=64%, age=64±30 yrs.), were measured. One patient data was removed. The mean BCM HYD status was 1.86 l, SD 1.46 l, and SEM 0.22 l. Cella was 1.806 l, SD 1.36 l, and the SEM 0.21 l. The 95% difference confidence interval (CI) was -0.66 to 0.55 l. The Pearson’s correlation (r) was r^2 = 0.85 (p<0.00001). There was no proportional bias: the offset was -0.056 l, and K=1.010. The limits of agreement (LOA) analysis showed a mean difference of 0.56 l, and limits d ±2SD = (−1.192 l, 1.081 l), indicating 95% of the difference will lie within these limits. To evaluate equivalence, we performed two one-sided t-tests (TOST). When the bounds were reduced to the limit =0.47 l and -0.59 l, we obtained a 0.046 p-value (alpha =0.05), at 80% statistical power. For 26% of the subjects, the difference was <0.1 l, for 43% <0.25 l, for 71% <0.5 l, for 83% <0.75 l, for 90% <1.0 l, and for 9.5% (4 patients) more than 1 l. Only two cases (4.8%) were just over the ±2SD limit.

**Conclusion:** This study suggests the BCM and Cella devices can be used interchangeably.

## INTRODUCTION

Fluid overload (FO) and depletion (FD) measured by the FMC BCM, are strong predictors of mortality for kidney dialysis patients. ^1^ Several under powered randomized clinical trials (RCT) have also reported improved outcome when the BCM device is used to help guide clinical fluid management, and larger RCT’s are ongoing. A medical BIS device was first introduced in 1991 and became the basis for the FMC BCM in 2001. ^2-4^

The BCM is now used throughout the world and is the fluid assessment device used in most kidney dialysis studies. However, the FMC BCM is large and expensive. The Cella Medical company has developed a small low-cost wireless BIS device named Cella for larger scale use. The purpose of this study was to compare the equivalence of the predicted HYD status by the two devices.

## METHODS

In this prospective diagnostic equivalence study, we measured the HYD status of maintenance HD patients at the Slovenian Novo Mesto Hospital Dialysis Center. Measurements were taken at patients’ normal time and day as they presented for their dialysis therapy. The clinical and measurement guidelines and protocol recommended in the FMC BCM User’s Manual was followed. Following Slovenian National Ethics Commission approval for the study (doc number: 0120-493/2019/15), and receipt of patient written informed consent, the height and weight of 42 dialysis patients (M=64%, age=64±30 yrs.), were measured pre dialysis. Then they were also measured by both devices’ pre-dialysis. Patient inclusion and exclusion criteria, as well as measurement protocol was also followed according to the BCM instructions. The BCM is an EU CE Mark Class IIa device. Cella has been certified to meet all equivalent US FDA Class II safety and other required standards.

Patients were positioned supine in the dialysis recliner chair. On the same side of the body, two ECG type electrodes were placed on each hand-wrist/foot-ankle for current signal input & voltage detection, respectively (Figure 1). BCM signal & detection leads were then attached. Typically, the right side is measured but the non-fistula side is required. Measurements were first made with the BCM, then the BCM cable leads were disconnected from the electrodes and Cella leads were connected, and the second measurement was performed. The entire process of data input, electrode placement and cable connection take only a few minutes, but the measurement itself takes only several seconds for Cella, and a minute or so for the BCM.

**Figure 1.**
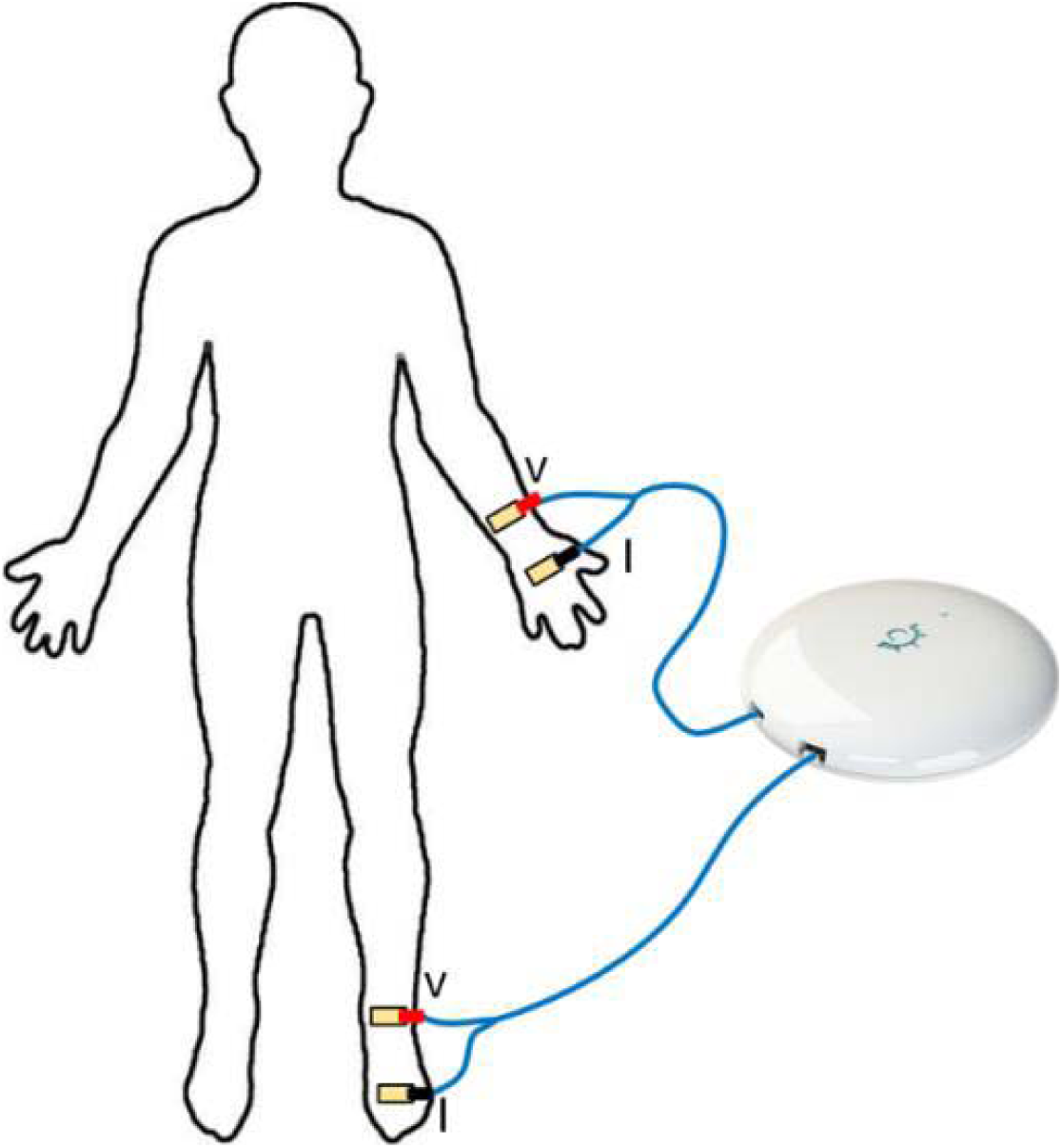
Measurement electrode and cable positioning. Typically, right side but non-fistula side required. The “I” represents current signal input electrodes and “V” is for voltage detection. Real Cella device size has 8cm diameter. From Cella Body Composition Analyzer User’s Manual: v1.04 2019, Cella Medical, Inc.

As described by FMC, the BCM estimates HYD using a three-compartment (3C) model whereby fixed intra and extracellular fluid (ICF and ECF) volume ratios expected for healthy normally hydrated lean and adipose tissues are compared to patients with end stage renal disease (ESRD), and any ECF above or below the 10^th^ and 90^th^ percentile (∼1 l) is considered FD or FO, respectively. ^5^

Cella Medical has developed entirely different patent pending methods to provide the equivalent information. ^6^ Using a large data set (3,000) of healthy normally hydrated subjects (US NHANES Survey), we computed HYD status by plotting ECF and ICF normalized to body weight (Wt). As the %ECF/Wt deviates from zero (regression line), HYD status can be computed in absolute liters (Figure 2). To express the Cella HYD result comparable to the relative BCM result, we normalized the absolute Cella HYD status to total ECF liters to obtain percent HYD. We also computed the 10^th^ and 90^th^ percentile range for normal hydration and found it to be ±9% (Figure 2). FMC reports it to be ±7%, which translates to ∼1.0 l for an average normally hydrated subject. Although HYD is thought to mostly be an ECF question, an abnormal relationship between ECF and ICF can be caused by change in ICF due to malnutrition. ^5^ Cella modeling supports an age correction based 3C (compartment) model. The age correction Cella uses is based on a regression model applied to experimental data. Analysis was performed with Octave version 4.2.2 and Graphpad.

**Figure 2.**
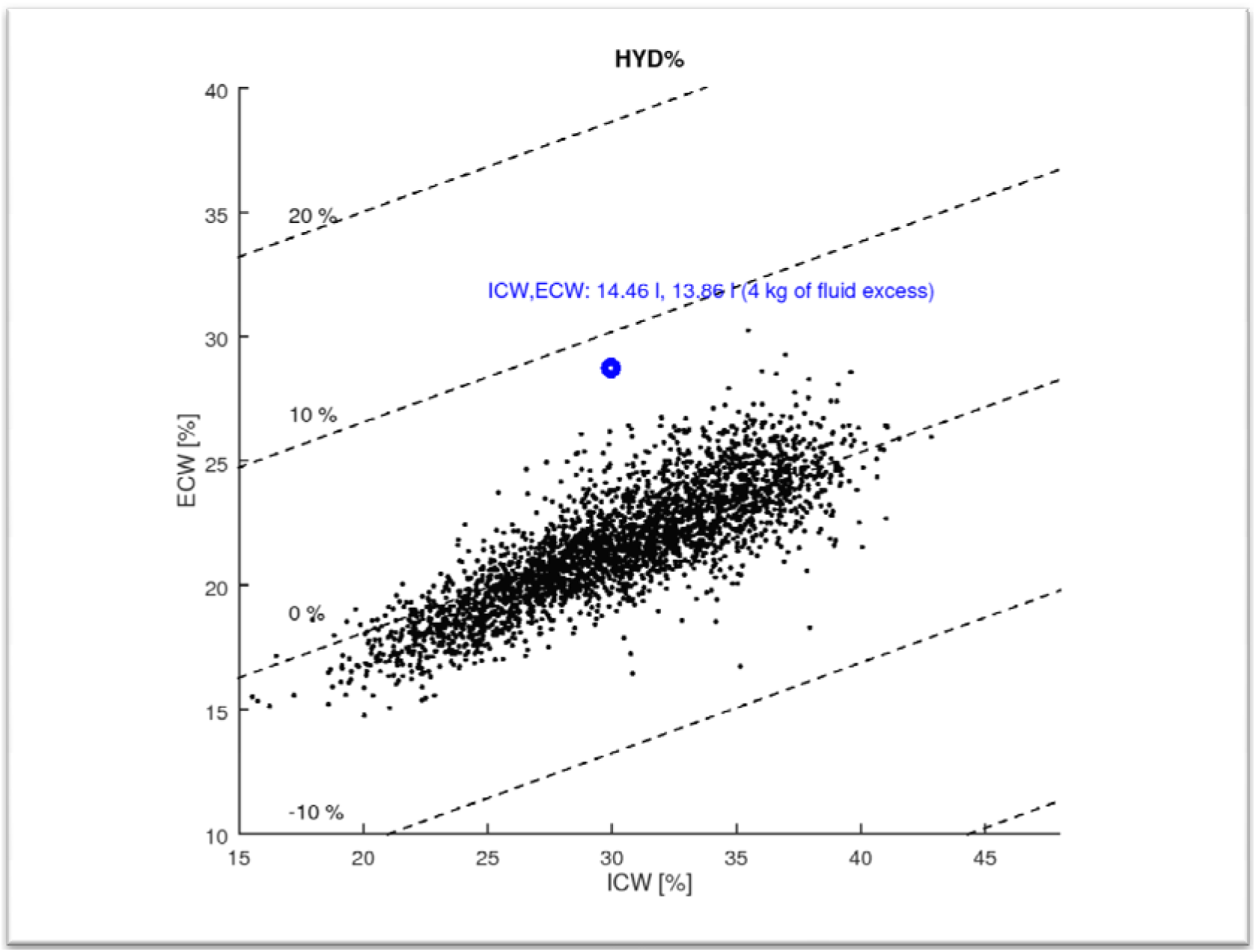
Cella Method of Determining Hydration (HYD) Status (patent pending) • The plot in **Figure 2** is an example showing ECW, ICW and HYD normalized to body weight and expressed in percentage (with respect to body weight). • The HYD result shown (fluid excess) are from calculating the Cella 3C model using the exact isotope data FMC reported as example of determining HYD status with their 3C Model (**Chamney 5**). The results are identical. • Although not shown in this plot, to be further compared to the FMC BCM, the 10th to 90th percentile range of HYD can be computed by normalizing HYD to total ECW, for Cella the variation of HYD is less than ±9%. The FMC BCM reports ±7%.

### Statistical analysis

The mean and SD for patient descriptive characteristics were computed as the mean, SD, SEM and confidence intervals (CI). We then compute a Pearson’s correlation along with an estimate of proportional bias. A Bland-Altman Limits of Agreement (LOA) analysis was performed to measure bias, the SD of the difference, limits of agreement d ±2SD, and CI. To evaluate equivalence, we performed two one-sided t-tests (TOST). This is a test based on a definition of an upper and lower equivalence bounds on which hypothesis testing is separately applied. We first evaluated the sample size needed for adequate statistical power (SP). ^7,8^

## RESULTS

One patient data was removed. Although positive that both devices provided similar poor results, the predicted FO in liters was unrealistic (∼10 l). The mean HYD status predicted by the BCM was 1.86 l, the SD 1.46 l, and the SEM 0.22 l. The predicted mean for Cella was 1.806 l, the SD 1.36 l, and the SEM 0.21 l. The 95% difference confidence interval (CI) was -0.66 to 0.55 l.

The Pearson’s correlation (r) between Cella and BCM HYD was r = 0.922 with an r^2 of 0.85, indicating a likely strong linear relationship (p<0.00001). Further, we found there to be no proportional bias: the offset was -0.056 l, and K=0.990, which is close to 1. (Figure 3).

**Figure 3.**
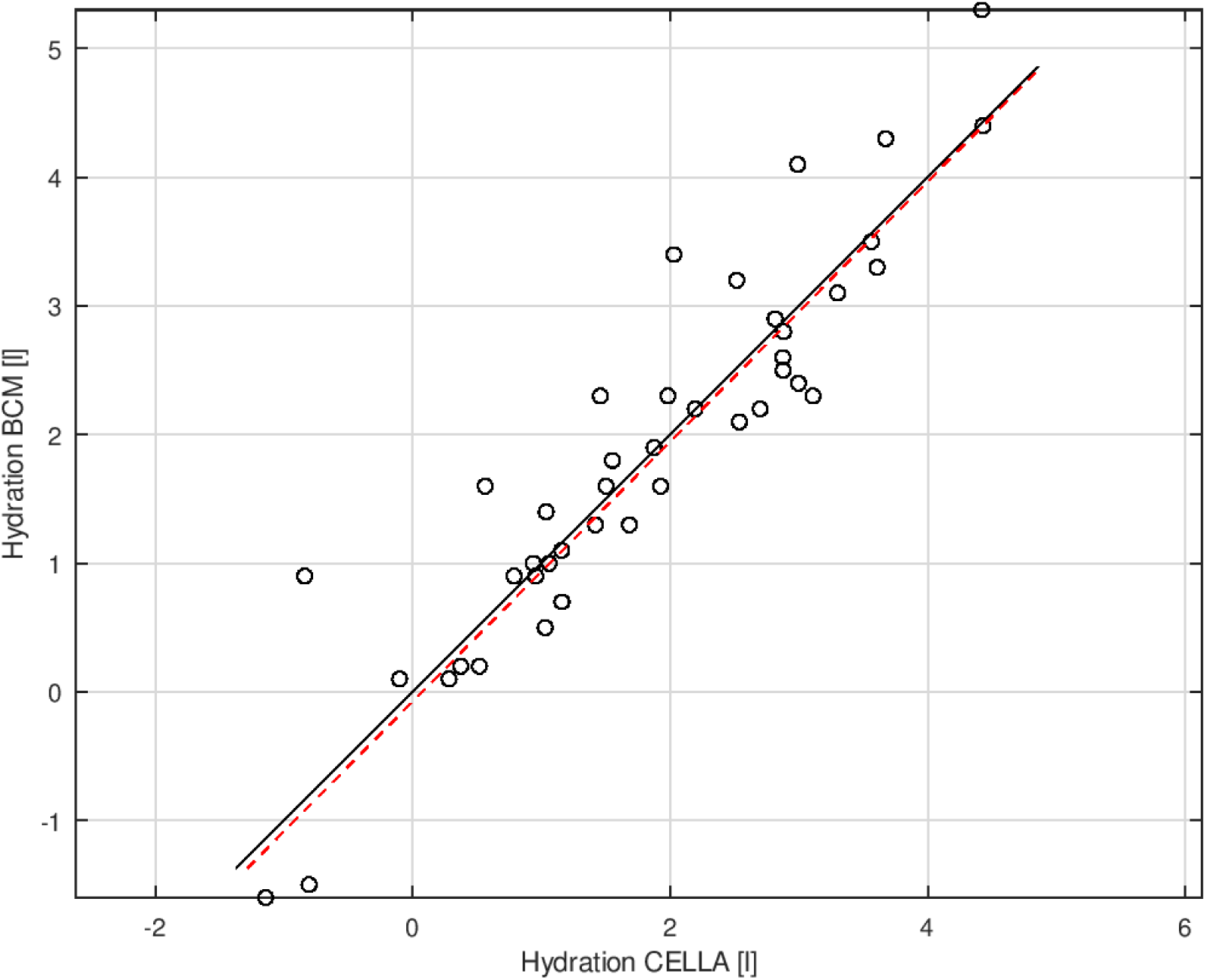
Correlation Analysis of Hydration Status. Measurements were carried out with BCM and CELLA on 42 subjects are plot and compared with line of equality (black). • The measurements are fit with a regression line (red): y=1.010 x – 0.074 • Correlation coefficient: R=0.9216, R^2^=0.8494 ∘ r=0.9216 (p<0.00001) For the null hypothesis that the measurements of the two methods are not linearly related. **Offset, proportional bias** • Offset means that two measured quantities x and y have a constant difference: e.g.: y=x+offset. The offset between Cella and BCM was -0.056 l, which is very small. • Proportional bias means that two measured quantities x and y are scaled the one with respect the other: y=x*K. Our analysis resulted in a K=1.010, which is very close to 1. So, there is no proportional bias between Cella and BCM.

Bland-Altman Limits of Agreement (LOA) analysis revealed a bias (mean difference) of only -0.056 l, a SD of the difference = 0.568 l, and a limits of agreement d ±2SD = (−1.192 l, 1.081 l), indicating 95% of the difference will lie within these limits. The precision of the LOA was SE 0.087 L for bias, LOA for SD was 0.152 l, and a CI: (−0.233 l, 0.122 l) thus 95% of cases the average comparative measurement would be in this interval. The lower limit 95% CI was: (−1.499 l, -0.885 l), and higher limit: (0.774 l, 1.388 l) (Figure 4).

**Figure 4.**
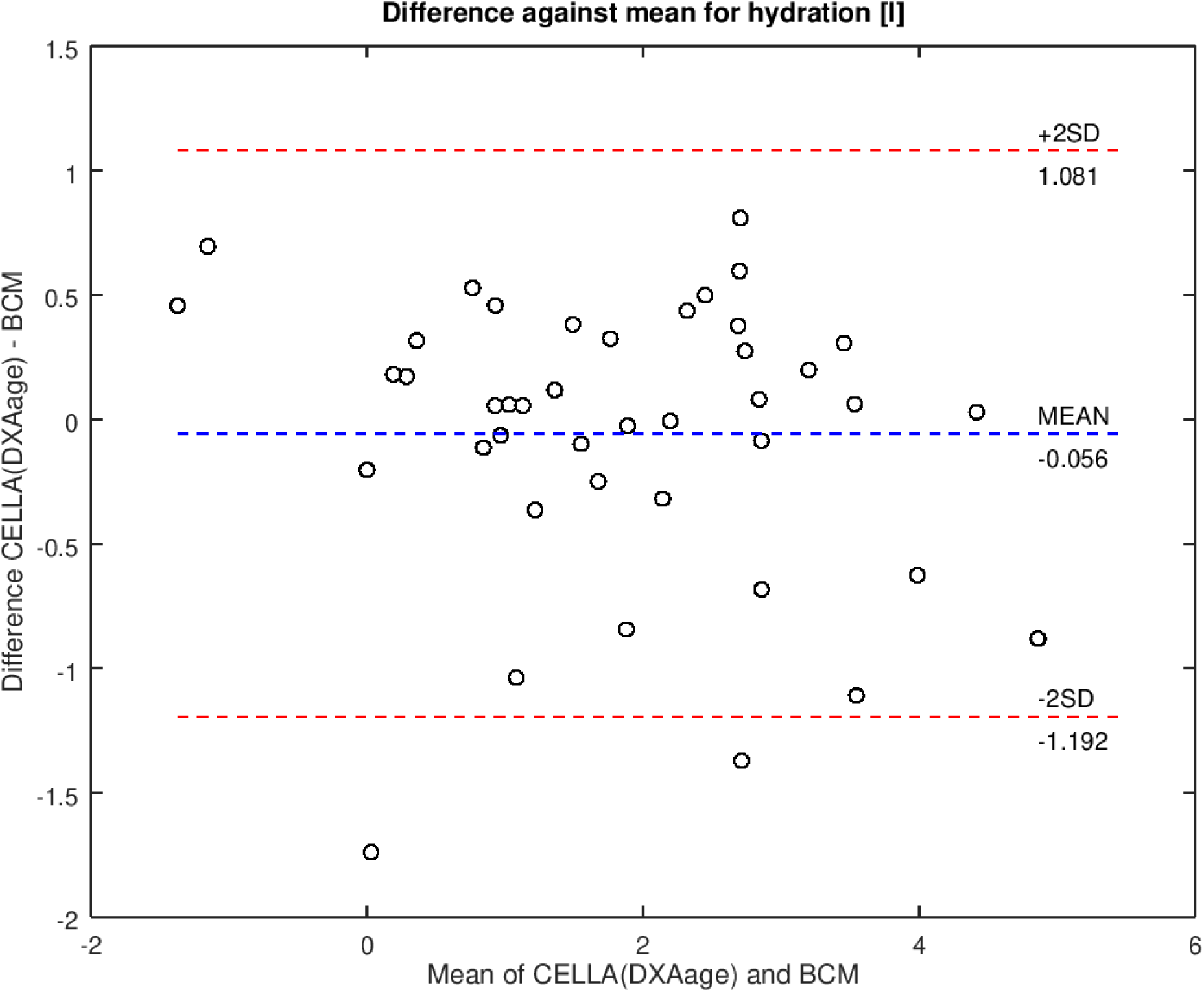
Analysis of Agreement. BIAS: d = -0.056 l (mean difference between CELLA and BCM) (Mean line) • Standard deviation: s= 0.568 l (standard deviation of the difference between CELLA and BCM) • Limits of agreement: d ±2s=[-1.192 l, 1.081 l] (interval where 95% of the difference will lie, ±2SD lines) **Precision of estimated limits of agreement** • Bias standard error: 0.087 l • Limits of agreement standard error: 0.152 l • Bias 95% confidence interval: [-0.233 l, 0.122 l] (That means that statistically 95% of the cases the average of comparative measurements between CELLA and BCM would be in this interval) • Lower limit 95% confidence interval: [-1.499 l, -0.885 l] • Higher limit 95% confidence interval: [0.774 l, 1.388 l] • The last two bullets mean that in 95% of the cases the limits of agreement will be according to the confidence intervals.

To evaluate equivalence, we performed two one-sided t-tests (TOST). This is a test based on a definition of an upper and lower equivalence bounds on which hypothesis testing is separately applied. We first evaluated the sample size needed for adequate statistical power (SP). For a difference of ±1 l, a sample size of n = 8.5 and n=10.4 required for 80% and 90% SP, respectfully. For ±0.6 l bounds, an n =23.7 and n=29.0 would be required for 80% and 90% SP. For +0.47 l and -0.59 l, an n=38.6 is required for 80% SP, and an n=47.27 for 90% SP. For hypothesis testing we report the highest result. The equivalence bounds (i.e., difference) for ±1 l, we obtained a p-value of 0.0015 (alpha =0.05), allowing rejection of differences larger than 1 l. For bounds ±0.6 l we obtained a p-value of 0.041 (alpha=0.05). If we further reduce the bounds to the limit =0.47 l and -0.59 l, we obtained a 0.046 p-value (alpha =0.05) at 80% SP. These results suggest the BCM and Cella results are equivalent.

For 26% of the subjects, the difference was <0.1 l, for 43% <0.25 l, for 71% <0.5 l, for 83% <0.75 l, for 90% <1.0 l, and for 9.5% (4 patients) more than 1 l. Only two cases (4.8%) were more than ±2SD outside the critical limit (Figure 5).

**Figure 5.**
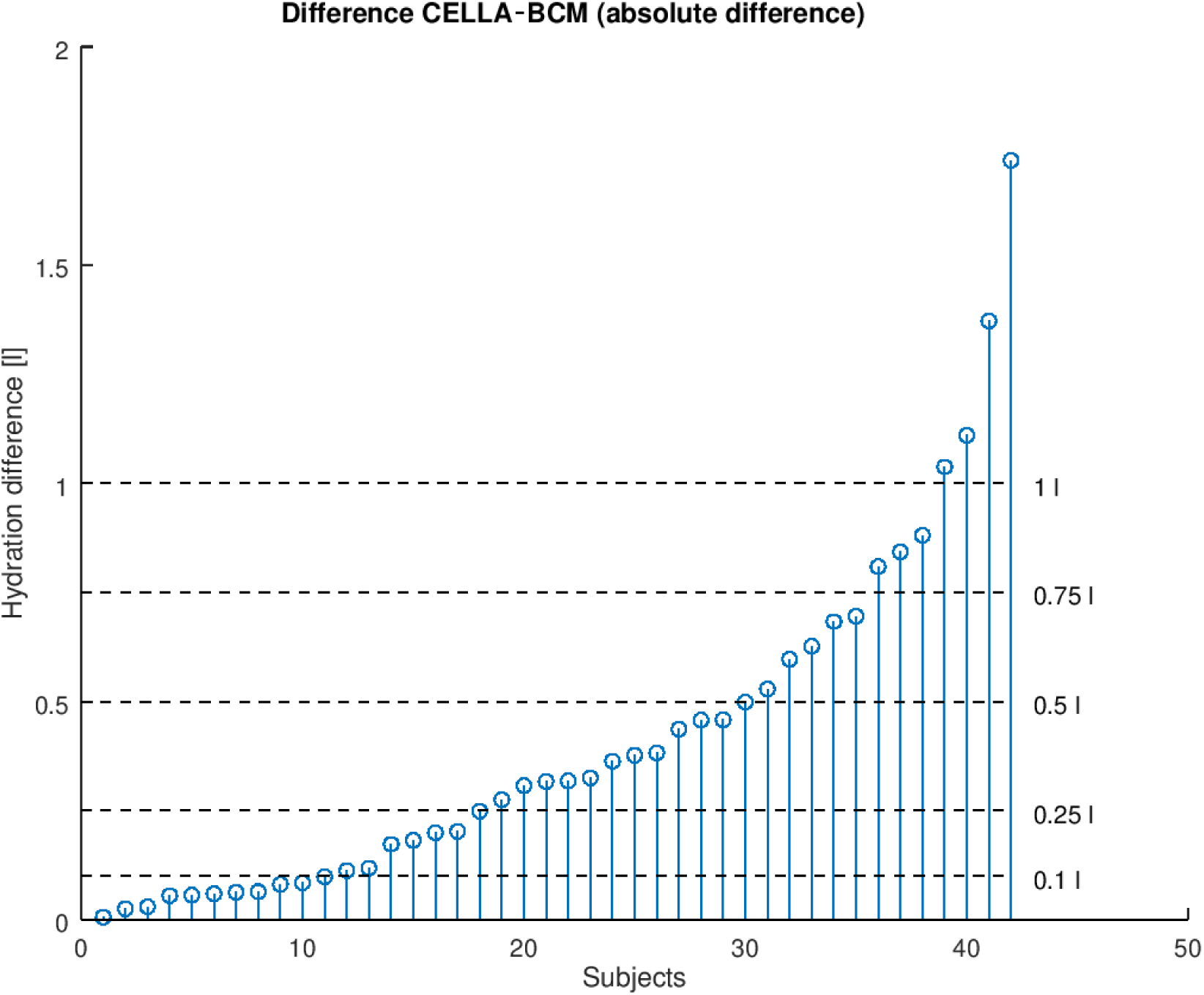
Hydration Difference Distribution. The comparative study between CELLA and BCM conducted on 42 subjects is showing: • 11 subjects with hydration difference < 0.1 l • 18 subjects with hydration difference < 0.25 l (about 43 % of the studied population) • 12 subjects with hydration difference between 0.25 l and 0.5 l (30 subjects less than 0.5 l) • 5 subjects between 0.5 l and 0.75l (35 less than 0.75 l, that is 83%) • 3 subjects between 0.75 l and 1 l (38 less than 1 l, that is 91 %) • 4 subjects above 1 l • 2 subjects outside the ±2SD range

Several interesting findings are revealed in Figure 6. But for one subject, the BCM and Cella results are all in in the same direction, so the differences are less. This also clearly indicates that the two methods are capturing the same phenomena. It is also important to note that the BCM consistently yields HYD results more extreme than Cella.

**Figure 6.**
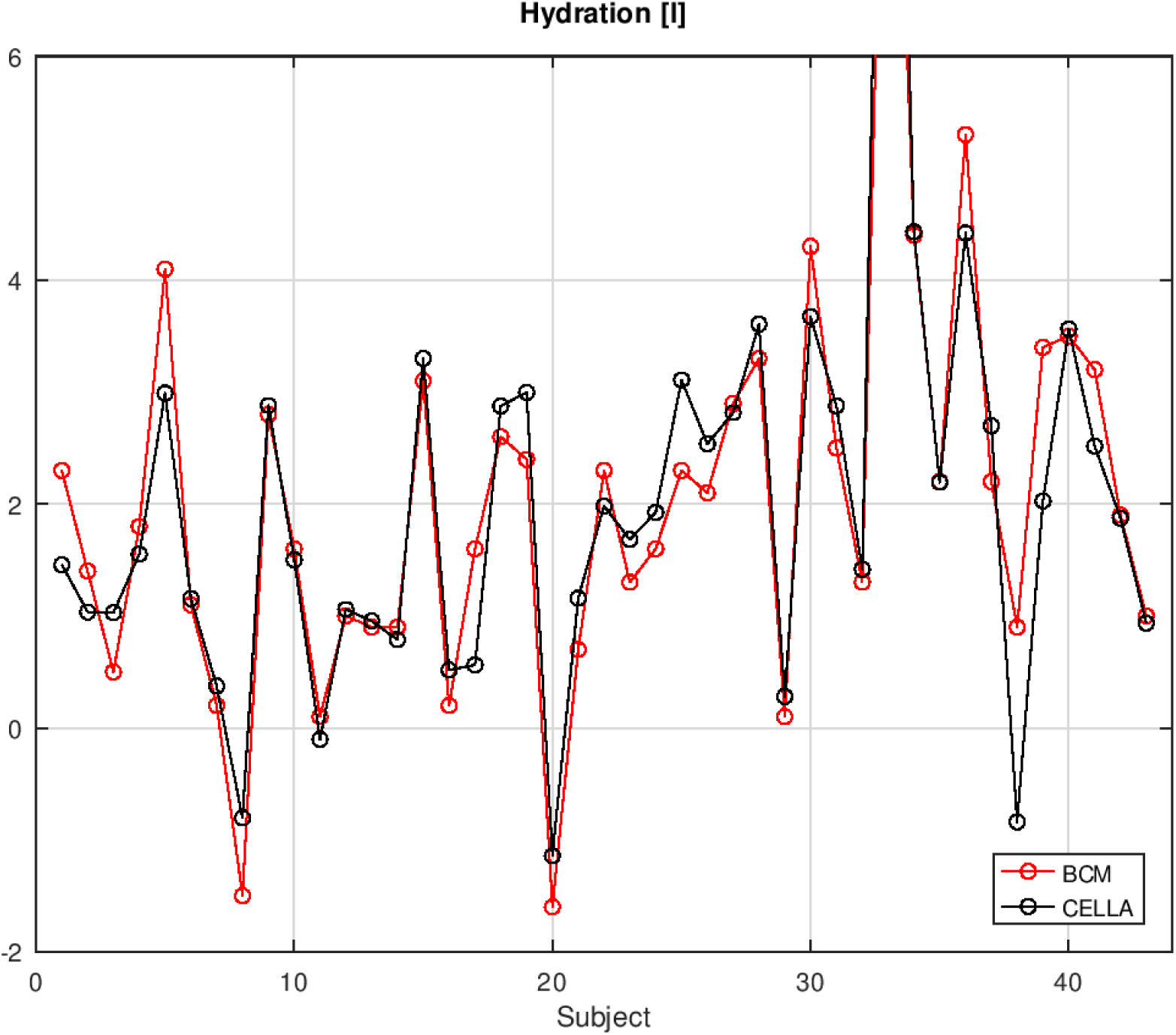
Comparison of Hydration Status (liters) Cella vs FMC BCM (43 dialysis patients) • Zero (0) represents the regression line through normally hydrated subjects in liters

## DISCUSSION

The results of this first small study are promising for people with ESRD. Furthermore, as the call for improving care and preventing chronic kidney disease (CKD) progression grows, the need for a low-cost high performance BIS device will increase. HYD needs to be better managed at home and in the clinic with more patient engagement. Overhydration has shown to be an independent predictor of CKD progression. ^9^

The FMC BCM represents a major turning point because it is was the first and only device that attempts to remove several major measurement error sources of estimating HYD status, allowing widespread use, acceptance, and progress. If the Cella device proves to provide results equivalent to the patented FMC BCM, ^10,11^ it would provide a lower cost alternative for larger scale use. There is presently no means of accurately predicting HYD status in liters other than with a BCM. Only by accounting for the water that would be contained in normally hydrated lean and adipose tissue, can HYD status be accurately determined. ^5^

The BCM has been a breakthrough device, providing important and unique clinical information that cannot be provided by any other methods. Thus, there is no way of directly validating the BCM HYD estimate. That the Cella device appears to obtain the equivalent results, using entirely different methods, in turn, provides validation for the BCM.

Of considerable relevance, the results of this study suggest the differences between Cella and the BCM may be important in a positive way. As shown in Figure 6, when there are larger differences between devices, the BCM seems to provide results more extreme (e.g., FO >4 l). A recent paper suggests that FO greater than 3 l would be large. ^12^ It would also seem extreme for FD to be -2 l. A recent RCT has reported a similar finding. ^13^ At a time when maintaining residual renal function is critical, a measurement of HYD volume erroring on the conservative side might be welcomed. ^14,15^.

## CONCLUSION

This study has found the Cella and BCM HYD results to be equivalent but further studies on larger more diverse samples are needed to replicate these findings.

## Supporting information

Equator Network Stard checklist

## Data Availability

As a non-interventional study, no data beyond what this paper reports are available. Replication of this study's results requires FMC BCM and CELLA BIS devices. Contact Cella Medical and Fresenius Medical Care for device availability.

## Author contributions

JM: study design, methodology, data interpretation, writing, review and editing manuscript. BB: statistical plan, software, data analysis, writing, review and editing manuscript. VMM: patient recruitment and coordination of measurements. BK: medical and ethics committee liaison, protocol, methodology, data collection, writing, review and editing manuscript. All authors contributed to the manuscript and approved the submitted version.

## Acknowledgments

We thank the patients for participating in the study and hope it will help improve their therapy.

